# High-intensity interval or circuit-based strength training effects on physical and cognitive functioning for community-dwelling older adults: A systematic review protocol

**DOI:** 10.1101/2022.02.10.22270669

**Authors:** Ashley Morgan, Kenneth S Noguchi, Ada Tang, Jennifer Heisz, Lehana Thabane, Julie Richardson

**Author notes:** Corresponding Author: Dr. Julie Richardson, School of Rehabilitation Sciences, McMaster University, Institute of Applied Health Sciences, 1400 Main Street West, Hamilton, Ontario, Canada, LSC 1C7.

## Abstract

**Background:** High-intensity interval or circuit-based strength training utilizes brief intense periods of strengthening exercises interspersed with rest/light activity or performed in a continuous circuit. The physical and cognitive effects of this time-efficient approach in community-dwelling older adults have not been systematically reviewed.

**Objectives:** To determine the effects of high-intensity interval or circuit-based strength training interventions on physical and cognitive functioning for community-dwelling older adults, and the associated adherence, retention, and adverse event rates.

**Methods:** Six databases will be searched from inception to November 2021; MEDLINE, EMBASE, CINAHL, AgeLine, SPORTDiscus, and Web of Science. To assess physical and/or cognitive functioning effects, randomized and quasi-randomized controlled trials (RCTs and Q-RCTs) of high-intensity interval or circuit-based strengthening interventions in community-dwelling older adults, utilizing any comparator, will be included. The above criteria will be extended to include non-randomized study designs to assess adherence, retention, and adverse events. All screening, data extraction and risk of bias will be assessed by two independent reviewers. Risk of bias will be assessed utilizing the Cochrane RoB 2 tool for randomized and ROBINS-I for non-randomized studies. Qualitative synthesis will be used to present information on adherence, retention, and adverse event rates. Qualitative and/or quantitative synthesis will be used to present physical and cognitive functioning effects depending on which method is deemed appropriate for the various outcomes.

**Conclusion:** This systematic review will be the first to summarize the physical and/or cognitive effects, adherence, retention, and adverse events reported in high-intensity interval and circuit-based strengthening interventions for community-dwelling older adults.

**Systematic review registration number:** CRD42021284010

## Introduction

Despite the recognized benefits of exercise on physical [1–5] and cognitive functioning [1,6–8] for older adults, only one third of older Canadians accumulate the recommended 150 minutes of moderate to vigorous physical activity per week [9]. The World Health Organization recommends older adults perform at least 150 minutes of moderate-intensity or 75 minutes vigorous-intensity aerobic activity per week or an equivalent combination, as well as the inclusion of strengthening exercises at least two days per week [10]. High-intensity interval or circuit-based strength training may offer a time-efficient form of exercise to address these recommendations.

Evidence suggests that exercise intensity may impact outcomes on physical [11] and cognitive functioning [12] in older adults. In older adults, recent systematic reviews found high-intensity aerobic activity demonstrates equivalent or superior effects on cardiovascular fitness compared with moderate intensity despite its’ shorter duration [11] and may facilitate greater improvements in working memory [12]. A systematic review of 56 meta-analyses of various exercise interventions for older adults reported that resistance training had the largest effect sizes overall, including for physical functioning [1], and other reviews suggest that higher intensity resistance training is superior to low intensity for improvements in strength [2,13] and aerobic capacity [2].

High-intensity interval training (HIIT), which combines brief intense bouts of aerobic activity combined with rest or low intensity exercise [14] is recognized as an efficient and effective mode of exercise with evidence supporting cardiovascular benefits and safety for older adults.

[11]. An analogous high-intensity interval or circuit-based format has also been applied for strengthening exercises [15–17], employing various descriptors; including, “high intensity interval in circuit training (HIICT)”[15] “high-intensity interval resistance training (HIRT)” and “high-intensity functional training (HIFT)” [18]. Through an interval or circuit format, strengthening exercises are performed at a high-intensity (relative to ability) with no or minimal rest, thereby also offering potential aerobic benefits [18]. Randomized controlled trials of high-intensity strength training using an interval or circuit-based format in older women have demonstrated improvements in walking speed [17] and speed of chair stands [16]. A 12-week quasi-randomized trial in older adults also found benefits on leg extensor strength, cardiorespiratory fitness (predicted maximal oxygen consumption [VO2max]) and reported all 18 participants in the high-intensity group completed the intervention with an overall attendance of 99% [19]. Despite the potential benefits, this type of exercise has received less attention than HIIT and the current evidence on the effects of high-intensity interval or circuit-based has not been systematically reviewed. The purpose of this review of this systematic review is to answer the following questions;

### Questions addressed by Systematic Review

1. What are the effects of high-intensity interval or circuit-based strength training interventions on physical and cognitive functioning for community-dwelling older adults? and,
2. What are the reported adherence, retention, and adverse event rates for these interventions?

## Materials and Methods

This protocol follows the Preferred Reporting Items for Systematic Reviews and Meta-Analysis Protocols (PRISMA-P)[20] (see Appendix I). This protocol is registered with the International prospective register of systematic reviews (PROSPERO), registration number; CRD42021284010.

### Inclusion criteria

#### Study Design

To determine effects of high-intensity interval or circuit-based strength training on physical and cognitive functioning, randomized controlled trials (RCTs) and quasi-randomized controlled trials (Q-RCT; allocation method not random e.g., alternation, day of week, etc.,) [21] will be included. To assess the reported adherence, retention, and adverse event rates, other non-randomized study designs, specifically nonrandomized controlled trial (NRCT), controlled before-and-after (CBA), and before and after (BA) studies [21] will also be included. Nonrandomized studies are prone to greater bias than RCTs but can be particularly useful in providing information regarding harms, especially when evidence from RCTs may be limited [22–24]. Grey literature will not be included.

#### Participants

Eligible participants will be community-dwelling older adults. Study samples must have a mean or median age of 60 years or older. Studies focused on samples of older adults with moderate/severe cognitive impairment (clinical diagnosis of dementia or Alzheimer’s disease or indicated by cognitive screening tool such as Montreal Cognitive Assessment or Mini Mental State Exam), a neurological condition or pre- or post-surgery will be excluded. Studies with mixed populations of older adults will be excluded if > 50% of the sample includes individuals with moderate/severe cognitive impairment (see previous criteria previously), a neurological condition or are pre- or post-surgery.

#### Interventions

Eligible interventions will be ≥ 4 weeks in total duration. Eligible interventions will involve strengthening exercises (including body weight and external resistance) that are performed in a continuous circuit (i.e., several exercises performed sequentially with no/minimal rest between exercises, may have rest between circuits)[25–27] or interval format (alternating periods of time performing high intensity exercise with periods of low intensity activity or rest[28]). High-intensity must be defined using prespecified target criteria (e.g., rating of perceived exertion, percentage of heart rate or one-repetition maximum). Acceptable criteria may include; ≥ 80% peak/maximum heart rate (PHR/MHR)[29], heart rate reserve (HHR)[30] peak work rate (PWR)[29], or maximal oxygen consumption (VO2 max)[30]; ≥ 70% one-repetition maximum (1RM); or ≥ 14/20 using Borg’s rating of perceived exertion scale [31,32] (or the equivalent of at least “hard” using another scale). Inclusion of a familiarization period (not meeting high-intensity criteria) is acceptable; however, this duration should not exceed 1/3 of the total intervention duration. Interventions which combine high-intensity interval or circuit-based strength training with other additional interventions likely to impact physical and/or cognitive functioning (e.g., cognitive training, other forms of exercise) will be excluded. ‘Other forms of exercise’ will be considered the inclusion of exercise components beyond a warm-up and cool-down period (e.g., high-intensity strength circuit followed by 30 minutes stationary cycling).

#### Comparators

To assess effects on the intervention on physical and/or cognitive functioning, studies comparing interval or circuit-based high-intensity strength training to a control group or intervention that does not include interval or circuit-based high-intensity strength training will be included. To assess adherence, retention, and adverse event rates, studies utilizing any or no comparator intervention will be included.

#### Outcomes

Eligible studies will be included if effects of the intervention on any specified physical or cognitive functioning outcome are included. Physical functioning outcomes will include measures of both physiological impairment; specifically measures assessing muscle strength or cardiorespiratory fitness, and measures of mobility and performance capacity [33] (e.g., self-report questionnaires, performance of functional tasks including walking and sit to stands). Cognitive functioning outcomes will include any measures which assess general cognition or specific domains/functions. Studies which only report acute effects (immediately after a single bout of exercise) will be excluded.

#### Setting

Any setting will be included provided the older adults are living in the community.

#### Language

Studies published in English will be included.

### Search strategy

Six databases will be searched with no date restrictions to October 2021: Ovid MEDLINE, Ovid EMBASE, EBSCOhost CINAHL, EBSCOhost AgeLine, EBSCOhost SPORTDiscus, and Clarivate Web of Science. A McMaster University research librarian assisted in the development and refinement of the MEDLINE search strategy which can be found in Appendix II. This strategy with feedback from the research librarian was adapted for each database listed above. Searches will be re-run in all databases in November 2021 to identify any additional studies. Finally, the references list of relevant studies will be hand-searched to identify additional studies.

### Data Management

Titles and abstracts of eligible articles will be uploaded to Covidence systematic review management software [34] for screening. After initial title and abstract screening, full text for eligible articles will be retrieved and uploaded into Covidence to assess against inclusion and exclusion criteria. The final data extraction phase will utilize Microsoft Excel software [35].

### Study Selection

Detailed inclusion and exclusion criteria will be provided to reviewers, titles and abstracts of articles will be screened independently by two reviewers (AM and KN), after a calibration period. This period will involve both reviewers independently screening 20 titles and abstracts to ensure inclusion and exclusion is understood. Articles that appear to meet inclusion criteria or are uncertain will be included at this stage. These articles will be retrieved in full text and will be screened independently by two reviewers (AM and KN) after a similar calibration period. Any disagreement will be resolved through discussion or consultation with a third reviewer (JR). Additional information will be sought from authors as needed. Reviewers will not be blind to study title or author(s).

### Data extraction

Piloted standardized data extraction forms will be used to extract data from each eligible article by two independent reviewers (AM and KSN). The data extraction forms found in Appendix III detail the variables for which data will be sought and these forms will be pilot tested with at least five articles to ensure completeness and clarity.

### Risk of Bias Assessment

Risk of bias for RCTs will be assessed using the revised Cochrane Risk of Bias (RoB 2) [36] tool. Non-randomized studies will be assessed using the Risk Of Bias in Non-randomised Studies-of Interventions (ROBINS-I) [37]. Two independent reviewers (AM and KSN) will conduct risk of bias assessments, with any disagreements resolved through discussion or consultation with a third reviewer (JR). Reviewers will not be blinded to article title or author(s).

### Data synthesis

The study characteristics reported in included studies will be summarized and compared to determine where synthesis may be appropriate. Study authors will be contacted to obtained insufficient or missing information. Information on the effects of the intervention on physical and cognitive outcomes will be collected from RCTs and quasi-RCTs. Information on adherence, retention, and adverse events will also be collected from the additional non-randomized designs specified in the inclusion criteria. Randomized controlled trials will not be combined with nonrandomized study designs for synthesis. Qualitative synthesis, utilizing text and tables, will be used to present information on adherence, retention, and adverse event rates and intervention characteristics (e.g., exercise parameters, duration). Data on the effects on physical and cognitive functioning outcomes will be grouped according to shared outcomes (e.g., muscle strength, gait speed, general cognition, etc.,) and presented using qualitative and/or quantitative synthesis depending on which method is deemed appropriate for the various outcomes.

Quantitative synthesis (meta-analysis) will be conducted based on considerations of clinical, methodological, and statistical heterogeneity. Clinical heterogeneity considers variability in the intervention and comparators, population characteristics, outcomes and follow-up duration[38] while methodological heterogeneity considers study design, measurement tools, and risk of bias. Clinical and methodological heterogeneity will impact statistical heterogeneity (variability in intervention effects). For those studies deemed suitable to combine based on clinical and methodological considerations; statistical heterogeneity will be quantified using the chi squared and I^2^ tests. An I^2^ statistic of 50-90% may represent substantial, and 75-100% considerable heterogeneity [38]. Combined studies with an I^2^ statistic of ≥ 75% will not be combined and those with 50-75% will consider the size and direction and effects and the strength of the evidence for heterogeneity (e.g., confidence interval for I^2^, p-value from chi-squared test) [38].

If appropriate, meta-analysis will be conducted using the mean difference for outcomes utilizing the same scale and standardized mean difference for those utilizing different scales [38]. Results will be presented with 95% confidence intervals. Missing standard deviation values will be estimated using standard errors, confidence intervals, t values, and/or P values based on the recommendations from the Cochrane handbook [38]. Data will be used from intention-to-treat analysis data if available. A random effects model will be used for any meta-analyses. If deemed appropriate for combination, subgroup analyses will be conducted based on intervention (criteria utilized for “high-intensity”; duration <12 weeks, ≥12 weeks) and sample characteristics (clinical conditions, sex). Credibility of subgroup analyses will be assessed using the Instrument to assess the Credibility of Effect Modification Analyses (ICEMAN)[39]. Sensitivity analysis will be conducted removing studies with high risk of bias.

### Assessing certainty of findings

The certainty of findings for intervention effects on physical and cognitive outcomes will be assessed using the Grading of Recommendations Assessment, Development and Evaluation (GRADE) [40] approach resulting a rating of high, moderate, low, or very low quality. GRADE summary of findings (SoF) tables will be used to summarize the results. The recommendations from the GRADE working group will be followed regarding the evaluation and incorporation of non-randomized studies into rating the certainty of a body of evidence[41], including rating the certainty of evidence for RCTs and quasi-RCTs separately.

## Discussion

High-intensity interval or circuit-based strength training may offer a time-efficient approach to exercise. However, existing evidence on the effects on community-dwelling older adults’ physical and cognitive functioning has yet to be systematically reviewed. Information summarizing the retention, adherence, and adverse events rates for interventions employing this type of training in older adult populations is also required. The results of this review will present valuable considerations for alternative training approaches and optimizing exercise prescription for community-dwelling older adults. High-intensity strength training using an interval or circuit-based format is an emerging area of research and this review will summarize the current evidence and recommend important future directions.

## Data Availability

All data produced in the present work are contained in the manuscript.

## Appendix I: PRISMA-P checklist

**PRISMA-P (Preferred Reporting Items for Systematic review and Meta-Analysis Protocols) 2015 checklist: recommended items to address in a systematic review protocol\***

| Section and topic | Item No | Checklist item |
| --- | --- | --- |
| **ADMINISTRATIVE INFORMATION** |  |  |
| Title: |  |  |
| Identification | 1a | Identify the report as a protocol of a systematic review<br>**Title and Abstract** |
| Update | 1b | If the protocol is for an update of a previous systematic review, identify as such |
| Registration | 2 | If registered, provide the name of the registry (such as PROSPERO) and registration number<br>**Abstract, PAGE 3** |
| Authors: |  |  |
| Contact | 3a | Provide name, institutional affiliation, e-mail address of all protocol authors; provide physical mailing address of corresponding author<br>**Title page** |
| Contributions | 3b | Describe contributions of protocol authors and identify the guarantor of the review |
| Amendments | 4 | If the protocol represents an amendment of a previously completed or published protocol, identify as such and list changes; otherwise, state plan for documenting important protocol amendments |
| Support: |  |  |
| Sources | 5a | Indicate sources of financial or other support for the review<br>**N/A** |
| Sponsor | 5b | Provide name for the review funder and/or sponsor |
| Role of sponsor or funder | 5c | Describe roles of funder(s), sponsor(s), and/or institution(s), if any, in developing the protocol |
| **INTRODUCTION** |  |  |
| Rationale | 6 | Describe the rationale for the review in the context of what is already known<br>**PAGE 1-2** |
| Objectives | 7 | Provide an explicit statement of the question(s) the review will address with reference to participants, interventions, comparators, and outcomes (PICO)<br>**PAGE 2** |
| **METHODS** |  |  |
| Eligibility criteria | 8 | Specify the study characteristics (such as PICO, study design, setting, time frame) and report characteristics (such as years considered, language, publication status) to be used as criteria for eligibility for the review<br>**PAGE 3-5** |
| Information sources | 9 | Describe all intended information sources (such as electronic databases, contact with study authors, trial registers or other grey literature sources) with planned dates of coverage<br>**PAGE 5-6** |
| Search strategy | 10 | Present draft of search strategy to be used for at least one electronic database, including planned limits, such that it could be repeated |

| **APPENDIX II** |  |  |
| --- | --- | --- |
| Study records: |  |  |
| Data management | 11a | Describe the mechanism(s) that will be used to manage records and data throughout the review<br>**PAGE 6** |
| Selection process | 11b | State the process that will be used for selecting studies (such as two independent reviewers) through each phase of the review (that is, screening, eligibility and inclusion in meta-analysis)<br>**PAGE 6** |
| Data collection process | 11c | Describe planned method of extracting data from reports (such as piloting forms, done independently, in duplicate), any processes for obtaining and confirming data from investigators<br>**PAGE 6** |
| Data items | 12 | List and define all variables for which data will be sought (such as PICO items, funding sources), any pre-planned data assumptions and simplifications<br>**APPENDIX III** |
| Outcomes and prioritization | 13 | List and define all outcomes for which data will be sought, including prioritization of main and additional outcomes, with rationale<br>**PAGE 7** |
| Risk of bias in individual studies | 14 | Describe anticipated methods for assessing risk of bias of individual studies, including whether this will be done at the outcome or study level, or both; state how this information will be used in data synthesis<br>**PAGE 6-7** |
| Data synthesis | 15a | Describe criteria under which study data will be quantitatively synthesised<br>**PAGE 7-8** |
| | 15b | If data are appropriate for quantitative synthesis, describe planned summary measures, methods of handling data and methods of combining data from studies, including any planned exploration of consistency (such as $I^2$, Kendall's $\tau$)<br>**PAGE 7-8** |
|  | 15c | Describe any proposed additional analyses (such as sensitivity or subgroup analyses, meta-regression)<br>**PAGE 7-8** |
|  | 15d | If quantitative synthesis is not appropriate, describe the type of summary planned<br>**PAGE 7-8** |
| Meta-bias(es) | 16 | Specify any planned assessment of meta-bias(es) (such as publication bias across studies, selective reporting within studies) |
| Confidence in cumulative evidence | 17 | Describe how the strength of the body of evidence will be assessed (such as GRADE)<br>**PAGE 8-9** |

## Appendix II: Search Strategy

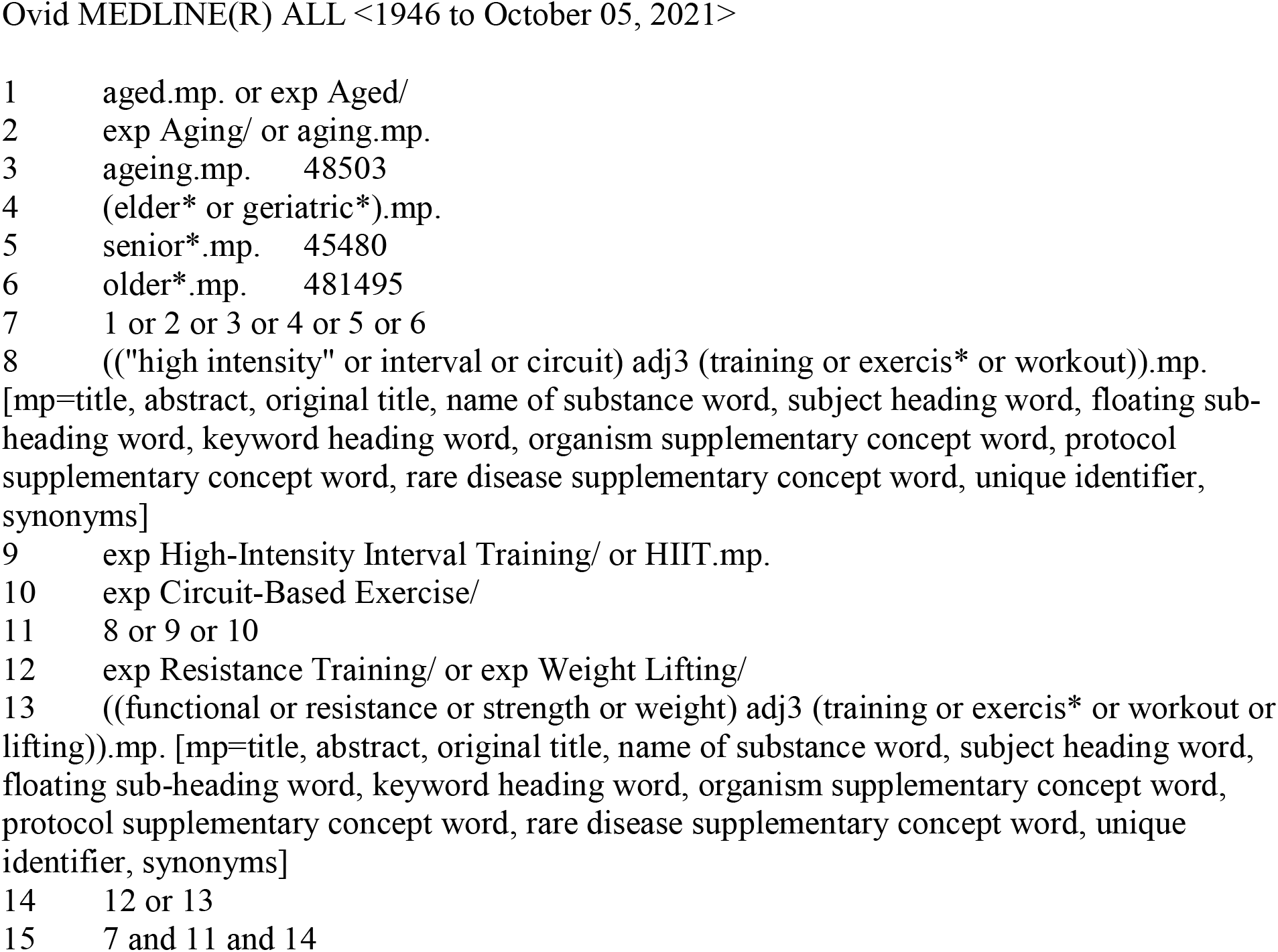

## Appendix III: Sample data extraction tables

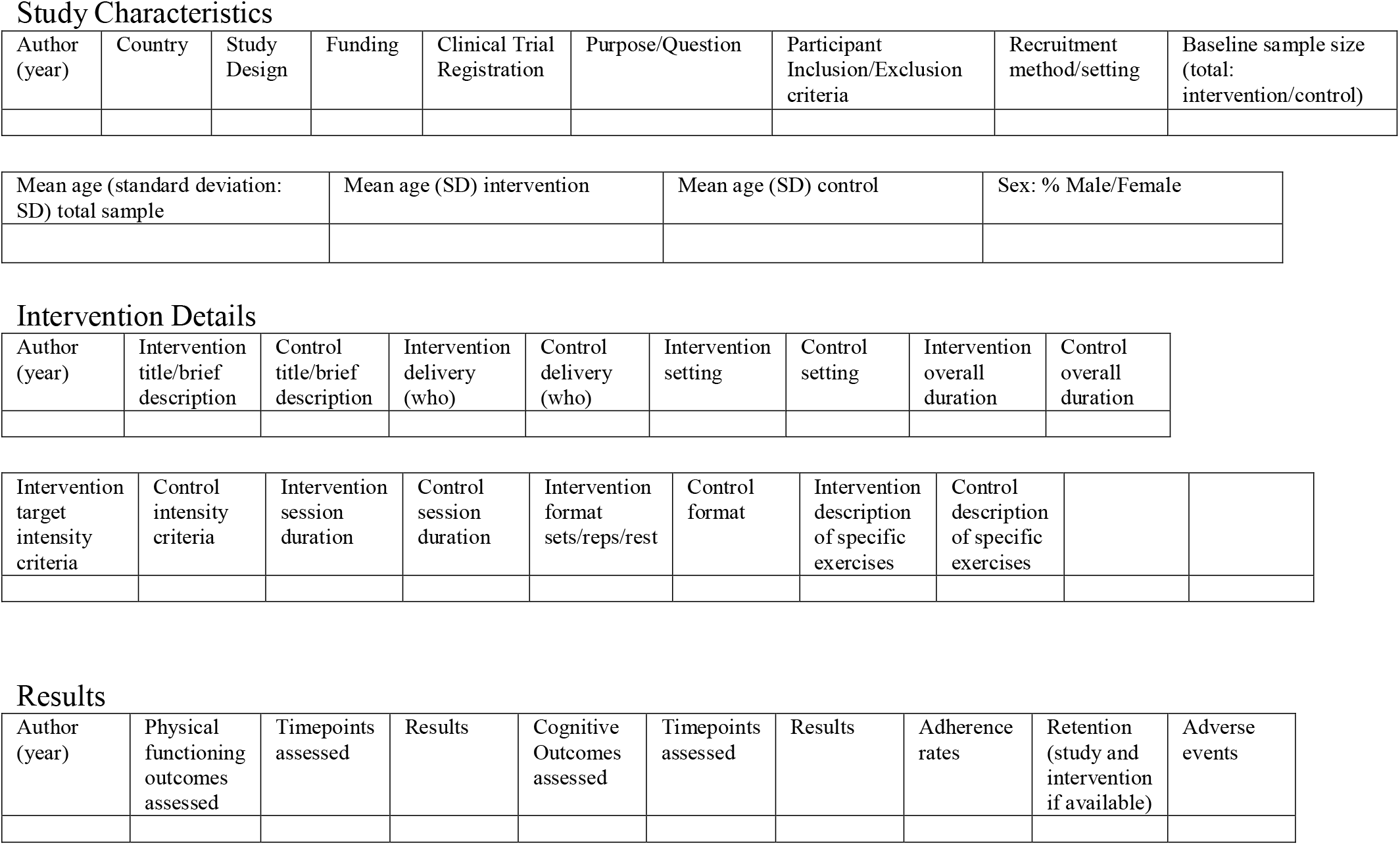

